# Social Capital Associates with Cognitive Health, Oral Health and Epigenetic Age Deceleration: A Cross-sectional Analysis of the Canadian Longitudinal Study on Aging (CLSA)

**DOI:** 10.1101/2023.07.06.23292314

**Authors:** A Liang, N Gomaa

## Abstract

**Objectives:** To quantify the association of social capital, defined as social relationships and networks, with cognitive health, oral inflammation, and epigenetic aging.

**Methods:** We used data from the Canadian Longitudinal Study on Aging (CLSA) (n=1,479; ages 45-85 years), categorizing social capital as structural and cognitive capital. Oral inflammation was determined as the presence of gum bleeding. Epigenetic aging was computed as the difference between chronological age and DNA methylation age. Multivariable regression models adjusted for covariates were used.

**Results:** Higher structural social capital was associated with decelerated epigenetic aging and better cognitive health outcomes. Higher cognitive social capital was also associated with better cognitive outcomes and less oral inflammation.

**Conclusion:** Enhancing social capital may contribute to better clinical and biological outcomes around aging.

**Visual Abstract:** 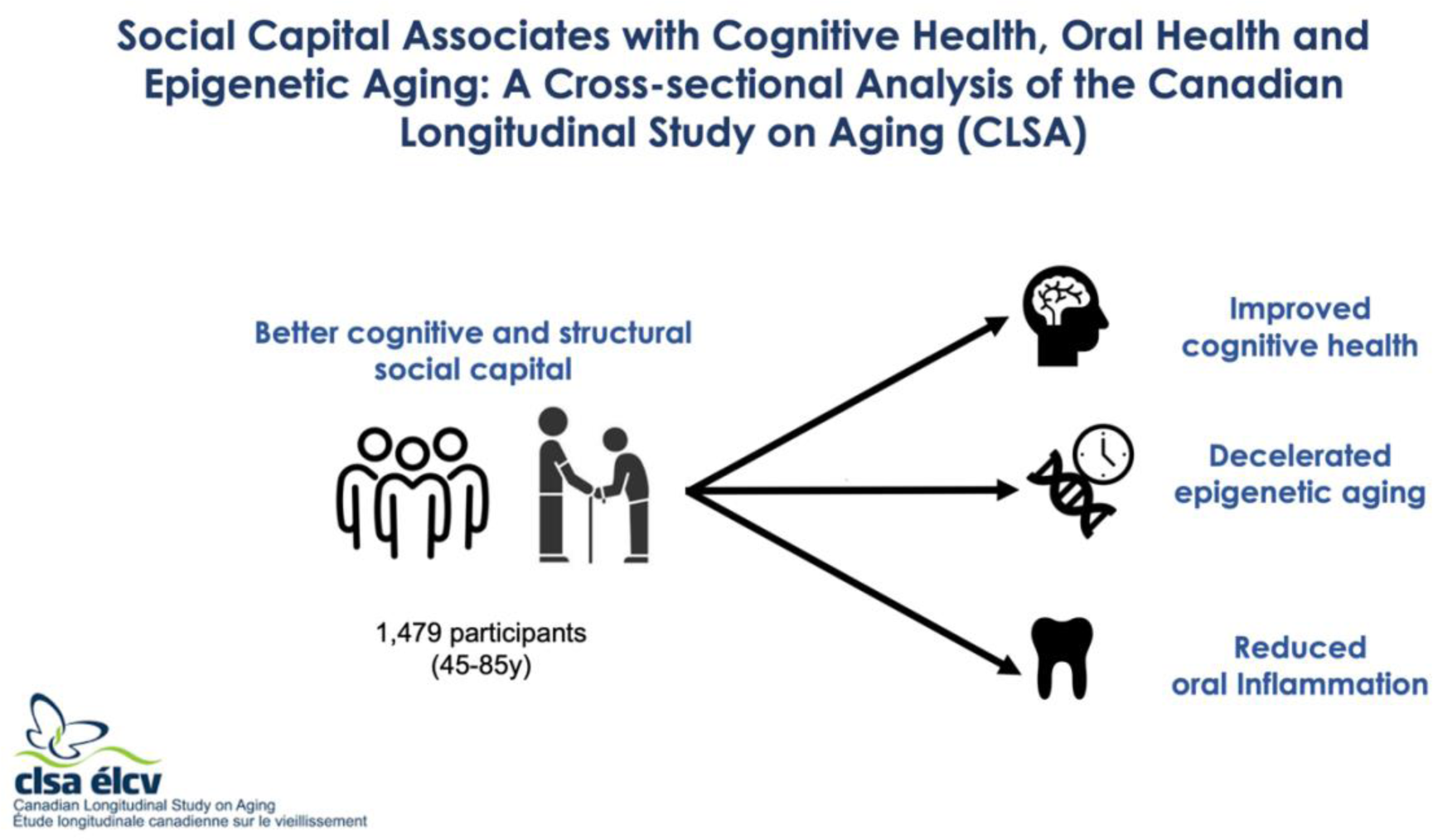

## 1. Introduction

Older adults are predicted to make up one third of Canada’s population in the coming decades (Hallman et al., 2022). Similarly, it is estimated that 1 in 6 people globally will be aged 60 and older by year 2030 (World Health Organization, 2022). Older age is a time in which a multitude of health conditions start manifesting and accumulating, placing a significant burden on individuals and societies. One health outcome that becomes of significant focus in older age is cognitive health. In Canada, the prevalence of mild cognitive impairments is estimated to be around 17% for adults over 65 years of age. For example, the prevalence of dementia doubles every five years from being less than 1% for those aged 65-69 to reaching over 25% for those above 85 years of age (Canadian Task Force on Preventive Health Care, 2015). Impaired cognitive health can have a profound negative impact on the quality of life, where performance in different cognitive domains including memory, attention, executive function, language, and psychomotor speed, can independently and collectively affect perceived physical and mental health, ability to complete instrumental and basic daily activities, as well as care required by caregivers and clinicians (Andrianopoulos et al., 2017).

Importantly, cognitive health has also been linked to several other health conditions such as metabolic health, cardiovascular health and oral inflammation. The link between cognitive health and oral health has been extensively investigated, mostly in association studies. Most of these have suggested oral inflammation, such as that caused by periodontitis to contribute to cognitive health decline due to the reduced sensory input related to the loss of masticatory contacts because of tooth pain or tooth loss (Dioguardi et al., 2020; Galindo-Moreno et al., 2022). As well, the oral cavity is the portal to the gut microbiome which has been implicated in cognitive health in various groups through the oral-gut-brain axis (Sansores-España et al., 2021). While several studies have assessed the mechanistic relationship between oral inflammation and cognitive health, it has been postulated that both outcomes are related through the common risk factors that affect individuals along the life-course and that can influence both groups of health conditions simultaneously from childhood to adulthood (Thomson & Barak, 2021). Some of these common risk factors are those that lie within the social and economic environment such as household income, one’s level of education and occupation and social class, with evidence consistently showing a gradient in most health outcomes by these social indicators (Gomaa et al., 2020). It is more plausible that the common social risk factors can elicit biological mechanisms that contribute to both outcomes rather than cognitive health and oral inflammation being linked biologically only. Despite the significant interest and debate around the common risk factors and their contribution to both cognitive and oral health, there have been sparse empirical evidence in support.

Social capital has gained significant attention as an important social determinant of health, especially around aging with studies showing it can play a major role in various health outcomes (Domènech-Abella et al., 2019; Ward et al., 2021). As described by Bordieu (1986) and Coleman (1988), social capital refers to the social resources that are available to both an individual and the community through their social relationships that are characterized by mutual trust, such as social networks (Bourdieu P., 1986; Coleman, 1988). Social capital can generally be categorized into cognitive and structural capital. Cognitive capital refers to subjective measures such as perceptions of social trust and social cohesion, while structural capital refers to objective measures such as social activity and social networks (Moore & Kawachi, 2017). Importantly, social capital has been emphasized as a predictor of various health outcomes in aging populations. Various studies have identified an association of social capital with all-cause mortality and self-reported general health (Fain et al., 2022; Verhaeghe & Tampubolon, 2012). As related to cognitive health, one cohort study showed an association between mid-life marital status and later cognitive functions, where being divorced or widowed was associated with greater odds of cognitive impairments later in life (Håkansson et al., 2009). Another cohort study showed social isolation and loneliness in older adults to be associated with lower cognitive scores (Lara et al., 2019). Likewise, oral health has also been found to associate with social capital, as have oral health related behaviours and access to dental care services (Campagnol et al., 2022; do Amaral Júnior et al., 2022). Recently, we have shown social capital to buffer the association between psychosocial stress and oral health in a large Canadian sample (Hensel & Gomaa, 2023).

Taking the above collectively, social capital can potentially be a common determinant for a plethora of health conditions including cognitive and oral health. This proposition has important implications to individual and population-level interventions which aim to target these common risk factors to alleviate multiple health conditions simultaneously, in other words the common risk factor approach to chronic disease prevention (Sheiham & Watt, 2000). Therefore, better understanding of the magnitude of the association between social capital and each of cognitive health and oral health is needed, with an ultimate goal of identifying targeted “upstream” interventions for the aging population that can improve health outcomes.

Meanwhile, common inflammatory mechanisms have been suggested to underlie several health conditions that relate to the aging process. *Inflammaging* is a relatively new term that was coined to describe the chronic, low-grade inflammation that may possibly underlie the pathogenesis of age-related diseases (Franceschi et al., 2018; Franceschi & Campisi, 2014). Inflammaging has been implicated in cognitive decline and chronic conditions such as cardiovascular, respiratory, metabolic disease and periodontal conditions, all of which associate with older age (Franceschi & Campisi, 2014). Previously, various biological indicators have been used to assess inflammaging including telomere attrition and shortening that contributed to cellular senescence that accumulates in different tissues, thereby contributing to age-related pathology (Franceschi & Campisi, 2014). Epigenetic-based biomarkers have also been strongly postulated to provide insights into inflammaging and to give an additional dimension to investigating the aging process. The most commonly investigated epigenetic marker in humans is DNA methylation (DNAm) which is the chemical addition of a methyl group to the cytosine residue, typically at cytosine–phosphate– guanosine (CpG) dinucleotides (Aristizabal et al., 2020). One approach to harness DNAm in relation to aging and thereby inflammaging is via the epigenetic clocks, which are a class of biological age estimators that utilize the level of DNAm at a set of computationally determined age-related CpG sites to estimate the biological epigenetic age (Aristizabal et al., 2020). The principal notion is to determine whether there are differences between the predicted epigenetic age and the actual chronological age of an individual (Hannum et al., 2013; Horvath, 2013). Such differences that reflect a variation in biological aging that can possibly be linked to experiences and environments on the one hand, or health outcomes on the other.

Clinical and epidemiological studies have found accelerated epigenetic age, where computationally estimated epigenetic age is “older” than chronological age, to predict mortality risk and associate with several health problems in adults such as frailty, cancer, cardiovascular disease, obesity, asthma, neurological diseases, and structural brain changes such as cortical thinning (Fransquet et al., 2019; Gomaa et al., 2022; Salameh et al., 2020). Moreover, observational studies have shown an altered epigenetic clock to be associated with higher risk of developing several neurodevelopmental and age-related conditions, as well as all-cause and cardiovascular mortality (Fransquet et al., 2019; Gomaa, 2022; Gomaa et al., 2022; Salameh et al., 2020). More recently, epigenetic age acceleration has been linked to adverse social and environmental conditions (Fiorito et al., 2017). Most studies on this front have identified links between epigenetic processes and socioeconomic status (SES) and/or psychosocial stress in various age groups. However, it is known whether social capital, or social relationships, may be related to the epigenetic aging process.

To this end, it is important that we identify the potential role of the common factors such as social capital that may contribute to health outcomes and potentially the biological mechanisms that can underlie the aging process such as epigenetic aging. In this study, we explore the associations between social capital and the outcomes that are of increased relevance to the aging population clinically and biologically, namely cognitive health, oral inflammation, and epigenetic aging. We assess these associations for each outcome independently with the hypothesis that enhanced social capital will be positively associated with better cognitive health, reduced oral inflammation and a deceleration of epigenetic age.

## 2. Methods

### 2.1. Data source and study population

In this cross-sectional study, we used data from the baseline wave of the Canadian Longitudinal Study on Aging (CLSA), which is a nationally representative longitudinal dataset that currently has data from over 51,000 participants collected via a combination of telephone surveys, at-home interviews, and data collection site visits.(Raina et al., 2009) Baseline data was collected in 2015 with ongoing follow-up data collection. Out of the total number of CLSA participants, our study comprised of the subsample of 1,479 participants aged 45 to 85 years, reflecting the total number of participants for which epigenetic data was collected and made available by the CLSA.

### 2.2 Variables

#### Independent variables

##### Social Capital

Two independent composite scores for structural and cognitive social capital were calculated. Structural social capital score was operationalized by summing the 20 questions from the CLSA focused on social participation. The questions relate to participation in community-related activities such as gatherings with friends, sporting events, and educational activities. Respondents receive one to five points based on how often they participate in each activity, with 100 being the maximum number of points possible on this continuous scale. Similarly, a composite score for cognitive social capital was summed from 19 questions on the CLSA based on the MOS-Social Support Survey, where participants respond on a five-point Likert scale. The total scores for structural and cognitive social capital were then standardized to a normal distribution with a mean of zero and a standard deviation of one. These were then converted to two independent categorical variables with every individual classified into having either high or low structural social capital and high or low cognitive social capital based on whether their scores were above or below the mean.

#### Dependent variables

##### Cognitive Health

Cognitive health was assessed as a continuous variable for each of the five domains of cognitive health: attention, verbal fluency, memory, executive function, psychomotor speed, which were obtained from participants’ scores using seven different cognitive assessments. Attention was assessed by the Stroop Neuropsychological Screening Test, which is scored by the time it takes to say the ink colour of each colour name. Verbal fluency was assessed by the average score of Animal Naming and Controlled Oral Word Association Test. The former is scored by the number of different animals recited in 60 seconds and the latter is scored by the total number of F, A, and S words recited in 60 seconds each. Memory was indicated by the average score of the Prospective Memory Test and the Rey Auditory Verbal Learning Test. The score for the Prospective Memory Test is derived from the participant’s intention to perform, accuracy of response, and need of reminders and the Rey Auditory Test is scored by the number of words correctly recalled in 90 seconds in the delayed recall. Executive function was assessed by the Mental Alternation Test, which is scored from the number of correct consecutive numeric and alphabetical alternations in 30 seconds. Lastly, psychomotor speed is calculated from the Simple and Choice Reaction Times, which assesses the mean reaction time of correct answers without outliers (Bayard et al., 2011; Teng, 1995; Troyer et al., 2006). The total scores for cognitive health in each domain were then standardized to a normal distribution with a mean of zero and a standard deviation of one, with higher numbers corresponding to better score.

#### Oral Inflammation

The CLSA assessed oral health using a seven-item questionnaire that gauges subjective measures of oral health by asking about participants’ experience with various oral health issues. Out of these questions, we selected the 3 questions that were used as proxy measures of oral inflammatory conditions. These included whether a respondent had any of the following conditions: gum bleeding, gum soreness, or burning sensation in mouth. Oral inflammation was dichotomized into a binary variable based on whether respondents answered “yes” to any of these conditions which was counted as them reporting oral inflammation.

#### DNA methylation analysis

The method for profiling genome-wide DNA methylation in peripheral blood mononuclear cells (PBMCs) in the CLSA participants has been described in the CLSA Data Support Document (David et al., 2020). Briefly, the proportion of methylation on cytoside-guanine (CpG) nucleotide base pairs on the DNA extracted from the PBMCs was measured using the Illumina Infinium MethylationEPIC BeadChip microarrays referred to as the EPIC arrays (Illumina, CA, USA). The EPIC array quantitatively measures DNA methylation at 862,927 CpG sites and 2932 CHH sites across the genome. To obtain the DNA methylation data, genomic DNA were extracted from the frozen PBMC samples using QIAsymphony DSP DNA Kits (Qiagen, Hilden, GE), and bisulfite conversion using the EZ DNA Methylation kit (Zymo, CA, USA) was performed. The resulting bisulfite-converted DNA was processed on the EPIC arrays following manufacturer’s instructions. For Quality control purposes, these raw array data were preprocessed using the GenomeStudio software (Illumina, CA, USA), which transformed the raw methylation values into beta values. The beta values range from 0 to 1 and indicate the proportion of methylation at each CpG loci present in the sample.

#### Epigenetic Age Difference

CLSA includes DNA methylation from 1,479 participants, which makes up the sample for our current study. From their methylation raw data, estimates of epigenetic age are provided as per several epigenetic clocks. Our study uses the epigenetic age based on the Hannum epigenetic clock algorithm. Epigenetic age difference was also provided as the difference each participant’s chronological age and their epigenetic age.

#### Covariates

We selected the covariates in this analysis guided by the literature. These included age, sex assigned at birth, race/ethnicity (reported as White and non-White), self-reported general health, health behaviours (frequency of smoking and alcohol consumption), and SES as indicated by annual household income (Diener et al., 1985; Kessler et al., 2003). We selected annual household income as the indicator of SES to control for as it generally tends to be a SES dynamic indicator as opposed to the level of education which can remain static over most of adulthood and may not reflect changes related to aging as well as income does.

### 2.3 Statistical Analysis

First, we applied descriptive statistics. Chi-squared test was used to assess the differences between high and low structural and cognitive social capital across the study variables. Univariate regression models were used to assess the crude associations between exposure and each of the outcome variables. We then constructed multivariable generalized linear regression models to assess the association between the social capital variables (structural and cognitive) and the continuous outcome variables: five domains of cognitive health, and epigenetic age difference. We used multivariable logistic regression models to measure the association of the social capital variables (structural and cognitive) with the binary outcome variable of oral inflammation. All multivariable regression models were constructed such that the covariates were added in a block-wise method i.e., Model 1 was adjusted for age and sex assigned at birth; Model 2 was additionally adjusted for self-reported general health, and Model 3 was additionally adjusted for SES as indicated by annual household income and health behaviours. All statistical analysis was performed using the software R4.0.4. Results were reported as β-coefficients and 95% confidence intervals for each of continuous variables of cognitive health scales and epigenetic age, and as odds ratios and 95% confidence intervals for the binary oral inflammation variable.

## 3. Results

### 3.1 Study population characteristics

The results of this study are described according to the STROBE reporting guidelines (Elm et al., 2007). The study population included 1,479 CLSA respondents. The mean age of respondents in this sample was 63.14 years and there was an almost equal distribution between the biological sexes, where females constituted 50.51% of the study sample. Almost half of the respondents had low structural social capital (n=708, 48%). Meanwhile, a total of 664 respondents (45%) had low cognitive social capital (Table 1). Compared to participants in the low structural social capital group, those in the high structural social capital group had an overall significantly better self-reported general health, higher income, higher education, were more likely to be non-smokers, home-owners, and were married or in a common law relationship, and had higher scores in all five cognitive domains as well as a smaller epigenetic age difference. Similar differences were observed between participants in the high cognitive social capital group and their counterparts in the low cognitive social capital group.

**Table 1.** Characteristics of study population stratified by structural and cognitive social capital: CLSA, 2015

|  | High Structural<br>Social Capital | Low Structural<br>Social Capital |  | High Cognitive<br>Social Capital | Low Cognitive<br>Social Capital |  |
| --- | --- | --- | --- | --- | --- | --- |
|  | n=771 (52%) | n=708 (48%) | *p* | n=815 (55%) | n=664 (45%) | *p* |
| **Age** | 63.45 ± 10.36 | 62.80 ± 10.22 | <0.0001 | 62.25 ± 10.18 | 64.18 ± 10.32 | <0.0001 |
| **Sex** |  |  | <0.001 |  |  | 0.8455 |
| Female | 424 (55) | 323 (46) |  | 414 (51) | 332 (50) |  |
| Male | 347 (45) | 385 (54) |  | 401 (49) | 330 (50) |  |
| **Race** |  |  | 0.3867 |  |  | 0.04174 |
| White | 723 (94) | 658 (93) |  | 771 (95) | 608 (92) |  |
| Non-White | 48 (6) | 50 (7) |  | 44 (5) | 56 (8) |  |
| **Self-Reported General Health** |  |  | <0.0001 |  |  | <0.0001 |
| Excellent | 160 (21) | 101 (14) |  | 181 (22) | 80 (12) |  |
| Very Good | 344 (45) | 248 (35) |  | 344 (42) | 248 (38) |  |
| Good | 208 (27) | 257 (36) |  | 229 (28) | 235 (36) |  |
| Fair | 56 (7) | 81 (11) |  | 53 (7) | 83 (13) |  |
| Poor | 3 (0) | 18 (3) |  | 8 (1) | 13 (2) |  |
| **Income** |  |  | <0.0001 |  |  | <0.0001 |
| Less than \$20,000 | 30 (4) | 64 (10) | | 31 (4) | 63 (10) | |
| \$20,000 - \$50,000 | 155 (21) | 209 (31) | | 158 (20) | 204 (33) | |
| \$50,000 - \$100,000 | 237 (32) | 221 (33) | | 249 (32) | 209 (34) | |

### 3.2 Higher structural and cognitive social capital positively associates with cognitive health in older adults

As shown in Table 2, multivariable models adjusted for age and sex showed a statistically significant positive association between higher structural social capital and four out of the five cognitive health domains which included attention (β 0.21, 95% CI [0.12, 0.30]), verbal fluency (β 0.16, 95% CI [0.11, 0.24]), executive function (β 0.18, 95% CI [0.084, 0.29]), and memory (β 0.08, 95% CI [0.012, 0.17]). The associations of structural social capital with verbal fluency and attention remained significant after adjusting for general health status and income, whereas the associations of structural social capital and the domains of executive function and memory were attenuated. Similarly, higher cognitive social capital was associated with domains of cognitive health including memory (β 0.010, 95% CI [0.023, 0.18]), psychomotor speed (β 0.14, 95% CI [0.042, 0.24]), verbal fluency (β 0.095, 95% CI [0.028, 0.16]), although the magnitude of the associations was larger with structural social capital than cognitive social capital (Table 2). These associations were also attenuated in fully adjusted models.

### 3.3 Higher cognitive, but not structural social capital, contributes to reduced oral inflammation

To assess whether structural or cognitive social capital was associated with oral inflammation, we conducted multivariable logistic regression models, controlling for covariates. Higher cognitive social capital was inversely significantly associated with oral inflammation in crude models (PR=0.94 95%CI 0.91, 0.97) (Table 3). This association was partly attenuated in models adjusting for general health status and was for fully explained when adjusting for SES and health behviours in fully adjusted models. Meanwhile, no association was observed between structural social capital and oral inflammation in this sample (Table 3).

**Table 3.** Crude and adjusted regression coefficients of the association between social capital and each of oral inflammatory load and epigenetic age difference: CLSA, 2015

| **Structural Social Capital** |  |  |  |
| --- | --- | --- | --- |
|  | **Model 1<sup>a</sup>** | **Model 2<sup>b</sup>** | **Model 3<sup>c</sup>** |
|  | OR (95% CI) | OR (95% CI) | OR (95% CI) |
| **Oral Inflammation** | 0.94 (0.93, 1.02) | 0.95 (0.94, 1.26) | 0.99 (0.95, 1.03) |
| **Cognitive Social Capital** |  |  |  |
|  | **Model 1<sup>a</sup>** | **Model 2<sup>b</sup>** | **Model 3<sup>c</sup>** |
|  | OR (95% CI) | OR (95% CI) | OR (95% CI) |
| **Oral Inflammation** | 0.94 (0.91, 0.97)** | 0.96 (0.92, 0.99)* | 0.97 (0.92, 1.00) |
| **Structural Social Capital** |  |  |  |
|  | **Model 1<sup>a</sup>** | **Model 2<sup>b</sup>** | **Model 3<sup>c</sup>** |
| | $\beta$ (95% CI) | $\beta$ (95% CI) | $\beta$ (95% CI) |
| **Epigenetic age acceleration difference** | -0.80 (-1.5, -0.14)* | -0.72 (-1.4, -0.057)* | -0.79 (-1.5, -0.10)* |
| **Cognitive Social Capital** |  |  |  |
|  | **Model 1<sup>a</sup>** | **Model 2<sup>b</sup>** | **Model 3<sup>c</sup>** |
| | $\beta$ (95% CI) | $\beta$ (95% CI) | $\beta$ (95% CI) |
| **Epigenetic age acceleration difference** | -0.056 (-0.71, 0.60) | 0.060 (-0.59, 0.73) | 0.13 (-0.57, 0.82) |

a. Adjusted for age, sex and race/ethnicity.
b. Additionally adjusted for general health status.
c. Additionally adjusted for income and health behaviours

### 3.4 Social Capital and Epigenetic Age Acceleration Difference

To assess whether either type of social capital was associated with epigenetic age difference, we conducted multivariable linear regression models. The results showed a significant negative association between high structural social capital and epigenetic age difference in all three models (Table 3) (β -0.79, 95% CI [-1.5, -0.10]. Cognitive social capital on the other hand was not associated with epigenetic age acceleration difference (Table 3).

## 4. Discussion

In the present cross-sectional study, we used baseline data from the CLSA to investigate social capital as a common determinant of clinical and biological outcomes that become particularly prominent around aging. The specific outcomes of interest in this study were cognitive health, oral inflammation and epigenetic aging. Our main findings suggest that social capital associates with all three outcomes and that its contribution can vary by the type of social capital i.e., objective structural measures versus the more subjective cognitive indicators of social capital. More specifically, our results showed that structural social capital indicated as social participation and social networks was associated with cognitive health and an accelerated epigenetic aging, whereas cognitive social capital, measured as the availability of social support, was associated with these outcomes, although with less of a magnitude of the association. Oral inflammation on the other hand was more significantly associated with cognitive social capital. Moreover, our findings demonstrated that socioeconomic factors such as income as well as health behaviours played an explanatory role in the link between social capital and all of the outcomes in this study.

Our results demonstrating that high social capital associates with better cognitive health outcomes are largely in agreement with previous studies that emphasize the role of social networks and relationships in cognitive health (Franceschi et al., 2018; Franceschi & Campisi, 2014; Håkansson et al., 2009; Lara et al., 2019). Interestingly, in fully adjusted models, the association with the domains of attention and verbal fluency remained significant. This is in line with previous systematic analysis by Kelly et al. that noted that the impact of social relationships on cognitive functions was more relevant to some domains of cognitive health that were particularly more affected than others (Kelly et al., 2017). A similar cross-sectional study by Fu et al. also found association between social activities and cognitive health (Fu et al., 2018). Furthermore, our findings demonstrating that the association of social capital with cognitive health was fully explained by SES and health behaviours, highlight their essential role as determinants of health. Interestingly, cognitive social capital but not structural demonstrated an association with oral inflammation in crude models, however this association was attenuated in fully adjusted models. These results align with recent findings in the Canadian adult population demonstrating the role of social capital and SES in oral health (Hensel & Gomaa, 2023).

Structural social capital showed a protective inverse association with epigenetic aging, where higher structural social capital decreased the difference between chronological and epigenetic age indicating a decelerated epigenetic clock. This demonstrates that better social networks and relationships can contribute to protecting against biological aging. Thus far, numerous studies have shown that early-life social relationships and environment can impact DNA methylation patterns (M. Szyf, 2012; Moshe Szyf, 2011), suggesting a biopsychosocial mechanism where stressful exposures due to poor social relationships can contribute to disease-inducive DNAm patterns through the activation of the hypothalamic-pituitary adrenal axis and the subsequent biological stress response (Gomaa et al., 2019). We therefore provide here insight that these mechanisms may also be at play in the aging population, where social activities and relationships can also have a role in altering the epigenetic clock. Further investigations that consider these associations from a mechanistic perspective will be worth pursuing.

Importantly, our results contribute to the body of the literature showing the role of social relationships and networks in various outcomes in the aging population. In particular, we show that social capital may be a common determinant and a pivotal player in clinical and biological outcomes that are informative of the aging process.

There are several strengths to our current investigation. First, our use of CLSA which is a large, population-based dataset. Another strength of our study is our use of both types of social capital as structural and cognitive. While structural social capital assesses formal structured opportunities or activities for social participation, such as volunteering, group membership, sports, and churchgoing, cognitive social capital refers to one’s subjective feelings of social trust, cohesion, sense of belonging, and loneliness (Moore & Kawachi, 2017; Xue et al., 2020). Both measures are closely intertwined, but may or may not coexist in the same individuals and may vary in the way they impact health outcomes. Analyzing both types of social capital separately allowed us to assess the specifics of subjective and objective social capital that contributed to the health outcomes in study. There also a few limitations in this study. First, this study was a cross-sectional one and therefore no causal inferences or temporality could be established between measures of social capital and any of the outcomes assessed. Also, the included measures of oral inflammation were self-reported as CLSA does not clinically assess oral health. While self-reported oral health has generally been shown to reflect those observed through clinical assessments, the indicators of oral inflammation in the CLSA questionnaire may not be entirely specific to inflammatory conditions affecting the mouth. Meanwhile, it is important to note that while we aimed to assess the independent contribution of social capital to each of the outcomes assessed, the goal of this work was not to quantify any links between cognitive health and oral inflammation or to identify mechanistic associations with epigenetic aging.

Overall, our findings contribute to the growing body of knowledge that supports the need for interventions to close the social gap in health and yield promising results that will need further investigation to decipher the role of the epigenetic clock in general and oral health outcomes over time.

## Data Availability

Data are available from the Canadian Longitudinal Study on Aging (www.clsa-elcv.ca) for researchers who meet the criteria for access to de-identified CLSA data

## Acknowledgement

This research was made possible using the data/biospecimens collected by the Canadian Longitudinal Study on Aging (CLSA). Funding for the Canadian Longitudinal Study on Aging (CLSA) is provided by the Government of Canada through the Canadian Institutes of Health Research (CIHR) under grant reference: LSA 94473 and the Canada Foundation for Innovation. This research has been conducted using the CLSA dataset Comprehensive Baseline version 5.1 and Comprehensive Follow-up 1 version 3.0, under Application Number [2010027]. The CLSA is led by Drs. Parminder Raina, Christina Wolfson and Susan Kirkland. The opinions expressed in this manuscript are the author’s own and do not reflect the views of the Canadian Longitudinal Study on Aging.

## 7. Acknowledgment

This study is supported through grants to NG from the Schulich School of Medicine & Dentistry, Western University and Lawson Health Research Institute, London, ON, Canada. AL is supported through the 2020-2022 Summer Research Training Program (SRTP) at Schulich School of Medicine & Dentistry, Western University

